# Wartime food availability in the Gaza Strip, October 2023 to August 2024: a retrospective analysis

**DOI:** 10.1101/2024.10.21.24315753

**Authors:** Francesco Checchi, Mija-Tesse Ververs, Zeina Jamaluddine

## Abstract

**Background:** The Israeli operation in the Gaza Strip has damaged much of the territory’s food system, exposing vulnerable population groups to poor nutrition. We retrospectively estimated food availability per capita and the contribution of different food sources and categories during the first ten months of the war.

**Methods:** We described the composition and caloric value of food trucked into Gaza based on United Nations data over the period 7 October 2023 to 31 August 2024 and compared these with the Israeli government’s. We supplemented trucking data with information on baseline stocks, agricultural output, air or seaborne deliveries, population denominators and pre-war caloric intake estimates into a probabilistic simulation to estimate caloric availability in both northern and south-central governorates.

**Results:** Between October 2023 and April 2024, food trucks entering Gaza remained below pre-war levels. Israeli data offered higher estimates of food weight trucked in but appeared to feature extreme approximation. Following Israel’s takeover of crossings in May 2024, United Nations data likely featured underreporting, though even Israeli data indicated declining deliveries. The share of food categories changed little during the period analysed, but trucked-in food’s caloric value was lowest just as food was scarcest (February-March 2024). Trucks accounted for three-fourths of food in south-central Gaza, but <20% in the north; air and sea deliveries made up a small percentage. During at least 12 weeks in northern and 4 weeks in south-central Gaza, per-capita caloric availability was below the recommended intake.

**Conclusions:** Israel, as occupying power, did not ensure sufficient food availability throughout the analysis period, and its data appear unreliable. Existing stocks probably mitigated caloric deficits resulting from insufficient deliveries, but air and sea routes added little. Strengthened coordination of food deliveries may be warranted to optimise caloric quantity and dietary diversity despite aid restrictions.

## Introduction

The population of the Gaza Strip has experienced seven decades of protracted conflict and 16 years of enforced restrictions on trade and the movement of people and goods, including food [1]. Since the 7 October 2023 Hamas attacks, Israel has conducted large-scale aerial bombing and ground operations in Gaza, resulting in at least 41,272 deaths and approximately 1.9M people displaced as of end September 2024 [2].

Prior to the current war, 62% of Gaza’s households experienced food insecurity [3] and ≈1.7M people depended on United Nations (UN) humanitarian food and cash assistance [3], which covered up to 48% of poor households’ caloric intake [4]. Nutritional outcomes at population level reflected an energy-sufficient but insufficiently diverse diet [5, 6]. The burden of both acute and chronic malnutrition in 6 to 59 months old (mo) children was low (1% wasting, 9% stunting and 2% underweight [7, 8]) while adult (18 to 69yo) prevalence of obesity was high (28%) [9]. Of infants <6mo, 42% were exclusively breastfed [7] with high adoption of infant formula. Only 43% of children aged 6 to 23mo enjoyed minimum dietary diversity [7], anaemia (haemoglobin <11.5 g/dL) affected 25% of first-year schoolchildren [8] while 11% of households had a poor food consumption score [7–10].

The ongoing military operation has severely damaged or destroyed much of Gaza’s food system, including cropland area [11], the fishing fleet [12], flour mills [13] and bakeries [14–16], supply chains and food markets, with the consumer price index for food rising from 210 pre-war to 600 by March 2024 [17]. Simultaneously, Israel has placed enhanced restrictions on aid flows and distributions, closing all but two southern crossing points into Gaza up to May 2024 and rejecting multiple consignments for ostensible security reasons [18]. While most people in the North Gaza and Gaza City governorates had fled south by December 2023, those who remained appeared largely cut off from aid: the UN Relief and Works Agency for Palestine Refugees (UNRWA) last delivered food to the north on 23 January 2024 [19], being then barred from further deliveries [20], while the UN World Food Programme (WFP) ceased its food convoy operations to the north on 20 January [21], only resuming these on a limited basis in March. In December 2023 the Integrated Food Security Phase Classification (IPC), a multistakeholder initiative that conducts evidence and consensus-based analyses of acute food insecurity and malnutrition to inform global emergency response, classified 25% of the population in the northern governorates as experiencing catastrophic acute food insecurity [22], updating this projection to 55% in March 2024 [23].

Despite multiple signals of far-deteriorated food security, successive IPC reports acknowledge uncertainty regarding actual food availability over time in Gaza. Indeed, in March 2024 Oxfam claimed that the population in northern Gaza had only 245 Kcal per person-day available [24], while Israeli academics, working with data from the Israeli Ministry of Defence’s Coordination of Government Activities in the Territories (COGAT) agency, put this figure at 3160 for all of Gaza during January-April 2024 [25]. Since May 2024, the re-opening of crossings into northern Gaza and increased food deliveries appeared to mitigate food insecurity, though the IPC projected that 22% of Gaza would remain in catastrophic food insecurity conditions between June and September [26].

We retrospectively estimated food availability per capita and the contribution of different food sources and categories during the first ten months of the war (in north and central south Gaza). We compiled data on six food sources: (i) existing household stocks, (ii) humanitarian warehouses, (iii) private stores, (iv) agricultural production, (v) airdrops and maritime deliveries and (vi) trucked food aid. In a separate paper we will explore how caloric availability may have affected nutritional outcomes.

## Methods

### Study population and period

We considered the entire population of the Gaza Strip, stratified into a northern (North Gaza, Gaza City governorates) and south-central region (Deir al Balah, Khan Younis, Rafah). As of March 2024, an estimated 250,000 individuals remained in the north [27], from approximately 1,200,000 pre-war [28]. Our analysis spans 7 October 2023 to 31 August 2024. Gaza’s population, age and sex population distribution and expected proportion of pregnant and lactating women in 2023-2024 were obtained from UNFPA projections of the 2017 census [29]. The movement of population between each region during the war period was tracked over time based on monthly United Nations Office for Coordination of Humanitarian Affairs (OCHA) reports, with simple interpolation to connect available data points. We made no adjustment for excess deaths or out-migration.

### Data on truck deliveries

#### Alternative sources

From 7 October 2023 to 5 May 2024, UNRWA systematically monitored and published the composition of all trucks crossing into Gaza [30], which we downloaded. From 6 May 2024, following the Israeli Defence Force (IDF)’s takeover of the southern border crossings (hereafter ‘Rafah operation’), the UN could no longer collect comprehensive trucking data, though it continued to publish these; private/commercial freight became particularly challenging to obtain information on [30].

Separately, COGAT publishes a simpler dataset (https://gaza-aid-data.gov.il/main/) with breakdown by date, route (land, sea, air), category of consignment (‘food’ is one of these) and metric tons (mt) consigned. As COGAT data do not specify consignment contents Kcal from this source cannot be established.

Until Israel re-opened the northern Erez and Erez West crossings, trucks had to leave south-central Gaza to resupply the north. We reconstructed the number of these trucks over time based on published information and data shared by WFP. As no data on content were available, we simulated their caloric equivalent by repeatedly sampling from the empirical distribution of calories per truck obtained from the UNRWA dataset (see below and Figure S1, Annex). The remaining trucked food was attributed to the south-central region.

#### Management of the UNRWA dataset

After excluding five records with missing description of truck contents, the UNRWA dataset contained 35,379 records, 25,242 (71.3%) from the period before the Rafah operation. For 86.5% (29,611) records the number of pallets was provided in lieu of weight; pallets’ standard load was 637.5 Kg [23] and should not have exceeded 750 kg [31, 32]. We represented likely variability in actual pallet weight as a uniform distribution comprising ±20% of the standard load. After merging multiple records with the same truck ID, 35,330 individual truck records were identified. Most trucks carried a single item, but 5071 (14.3%) carried up to nine items. We allowed the relative weight share of each item, for multi-item trucks, to vary randomly. While there are reports of aid being rejected by the IDF even after a truck crossed into Gaza [18], we assumed that food would have been allowed through.

After harmonising alternative food item nomenclature, we applied each item’s caloric equivalent by consulting the NutVal 4.1 application (https://fscluster.org/sites/default/files/documents/nutval-4.1_0.xls) or the United States Department of Agriculture’s FoodData Central database [33]. Mixed or non-specific items were almost always sent by WFP, UNRWA, the Palestinian Red Crescent or World Central Kitchen (WCK) and its partner American Near East Refugee Aid (ANERA). We were able to decompose ‘food baskets’, ‘food cartons’, ‘food parcels’, ‘ration packs’ and ‘ready meals’ sent by the above organisations by sourcing their detailed basket/parcel compositions; we used the mean food category and caloric composition of the above organisations’ parcels to impute that of a minority of parcels sent by others. Similarly, for ‘food items’, ‘canned food’ and ‘cooked food’ we took the mean caloric value of individual (canned or any other) items specific to each of the above organisations, or the mean across all senders for other organisations. Lastly, specialised food items used to prevent or treat malnutrition (ready-to-use food, therapeutic milk, supplementary foods) were omitted from trucking data and instead sourced from the Nutrition Cluster dashboard [34].

### Data on other food sources

#### Existing household stocks of humanitarian food aid

Pre-war, UNRWA provided 1675 Kcal/day to 620,310 and 902 Kcal/day to 389,680 registered refugees in Gaza, distributed every three months [35], while WFP assisted 20% of non-refugee households (about 243,000 people) with food parcels [10]: households were classed into four vulnerability tiers, which received 204,000, 433,000, 837,000 and 1,054,000 Kcal every three months, respectively [36]; we assumed that an equal proportion of WFP-assisted households belonged to each tier. Altogether, humanitarian food assistance amounted to approximately 871 Kcal/day/person considering the whole population living in Gaza. We allocated this amount to the north and south-central regions based on the pre-war share of food aid recipients in each (49.4% and 50.6% respectively based on combined UNRWA and WFP data).

We assumed that at baseline UNRWA- and WFP-assisted households would have held between 0-100% of their last UNRWA or WFP rations, as detailed above, depending on the last distribution’s timing. We distributed this amount over the first three months of the crisis (namely the interval between distributions) but discounted it for the increasing percentage of damaged or destroyed residences in each region, as a linear interpolation of approximately monthly estimates provided by the United Nations Satellite Centre (UNOSAT) based on satellite imagery [37]: we assumed this percentage to be a proxy for household food loss.

#### Existing humanitarian warehouses

The exact content of food available in UNRWA warehouses on 7 October 2023 was shared by UNRWA. For WFP, we sourced data on existing warehouses’ location and storage capacity [38], which we transformed into a caloric equivalent based on the WFP ration’s contents; because warehouse capacity was given as a range (e.g. 100-500 MT), we sampled from this range during simulation (below). We assumed that all warehoused food became available, despite reports of Israeli airstrike damage and looting.

#### Existing private stores

Let *I*_0_ be the mean pre-war caloric intake per person-day (see below for its estimation). We assumed that *I*_0,*m*_, the fraction of *I*_0_ not met through food aid pre-war, would have been entirely sourced from private markets stocked from agricultural output within Gaza (see below) and retail food trucked in. By around 18 December 2023, the last of these private stores were reported to have emptied [39], suggesting that they had held ≤ 72 days of stock. Therefore, we assumed that stores would have been able to supply *I*_0,*m*_ for 72 days based on their baseline stock, but discounted this amount, as above, for the reported decreasing proportion of operational stores in each region over time [17, 39], which could reflect stockouts as well as military damage; as above, we linearly interpolated sequential data points of store operationality.

#### Agriculture and livestock production

Pre-war, agricultural output fulfilled about 12% of *I*_0_ [4], but remote sensing analysis showed 8% damage to cropland by December 2023 [40], 28% by January 2024 [41], 43% by February [42] and 57% by May [11]. We thus assumed roughly that the agricultural sector would have fulfilled 10-15% of *I*_0_ during the first two months of the crisis (uniformly distributed), with this proportion declining to 4-7% by May 2024 as a linear interpolation of percent cropland damage.

#### Airdrops and boat deliveries

Since late February 2024, countries including the United States, the United Arab Emirates, Egypt, Jordan, Germany, France and the United Kingdom carried out airdrops, mostly over northern Gaza. We reviewed press releases by each country’s armed forces to extract weight and caloric equivalents of each airdrop. Where caloric equivalent was not provided (9 instances), we assumed the same caloric equivalent as the WCK food parcels. Where medical supplies were reported to be part of the airdrop (5 instances), we assumed food accounted for 70% of total weight. We also included the boat delivery by WCK and partners on 15 March 2024 and all deliveries by the Joint Logistics Over-the-Shore (JLOTS) maritime corridor set up by the United States, which operated intermittently between 16 May and 17 July 2024 [43].

### Estimating per-capita caloric availability over time

#### Baseline and recommended intake

We computed the average recommended Kcal intake per person-day as the mean of recommended intakes [44] by age (0 to 4yo, 5 to 9yo, 10 to 14yo, 15 to 19yo, 20 to 59yo and ≥60yo) and sex, weighted by the proportion of Gaza’s population in each age-sex stratum, plus extra amounts required for pregnant and lactating women.

A self-weighting sample survey of non-communicable disease prevalence and risk factors among adults aged ≥40yo in Gaza was conducted in 2020 (ZJ was the lead survey analyst and HG the principal investigator) [6]. The survey collected individual characteristics including sex, age and diary-based Kcal/day intake. Of the 4,576 survey observations, 380 (8.3%) had missing values, leaving 4,196 in the analysis. We aggregated log caloric intake and its standard deviation by sex and age group (40 to 49yo, 50 to 59yo, 60 to 69yo, ≥70yo) and weighted each sex-age stratum estimate as above. We found that log intake featured a near-normal distribution (see Annex, Figure S2); we thus sampled random values from each age-sex stratum distribution and computed from these a weighted mean intake for persons aged ≥40yo. We imputed intake in younger age groups as the recommended intake times the ratio of actual (survey-estimated) to recommended intake among people ≥40yo. We took the resulting weighted average intake to be an estimate of the pre-war mean intake per person-day, namely *I*_0_.

#### Caloric balance and consumption during the war

To propagate uncertainty for each food source and parameter, we implemented 1000 runs of a simulation, during each run sampling randomly from distributions as described above and calculating net caloric balance at the end of each day in the analysis period as the difference between calories available and calories consumed. Calories available were the sum of remaining prevalent sources at the war’s start (warehouses, household stocks, private stores) and incident sources (agricultural output, trucked-in food, air and boat drops). We assumed that people in both northern and south-central Gaza would continue to consume *I*_0_ on average whenever sufficient food was available, or as much as possible if not. Accordingly, to estimate *I*_*r,t*_, i.e. the actual daily mean consumption in region *r* on day *t*, we estimated *K*_*r,t*_, the combined daily calories available from all the above food sources, and divided this by *N*_*r,t*_, estimated population at that time, constraining *I*_*r,t*_ ≤ *I*_0_. During each daily timestep, we depleted the starting caloric balance by *I*_*r,t*_*N*_*r,t*_. The quantity *I*_*r,t*_ is not technically an intake measure, as it does not account for food loss, but rather expresses the theoretical caloric availability per person assuming intake remained upper-bounded by *I*_0_. All analysis was done in R statistical software [45].

### Ethics and data availability

The study was approved by the Ethics Committee of the London School of Hygiene and Tropical Medicine (ref. 29926). All datasets (including URL sources for each datum) and R analysis scripts are available on https://github.com/francescochecchi/gaza_food_availability.

## Results

### Quantity of food trucked in

As shown in Figure 1, over the analysis period the number of food-transporting trucks that entered the Gaza Strip appeared consistently lower than the pre-war baseline of 150-180 per day. While the number of trucks rose steadily between late October 2023 and early January 2024, it then fell again, only reach pre-war levels in late April 2024. Since the Rafah operation in early May and its subsequent inability to consistently register truck consignments, UNRWA reported a consistently lower and declining number of trucks, albeit nearly all food-transporting.

**Figure 1.**
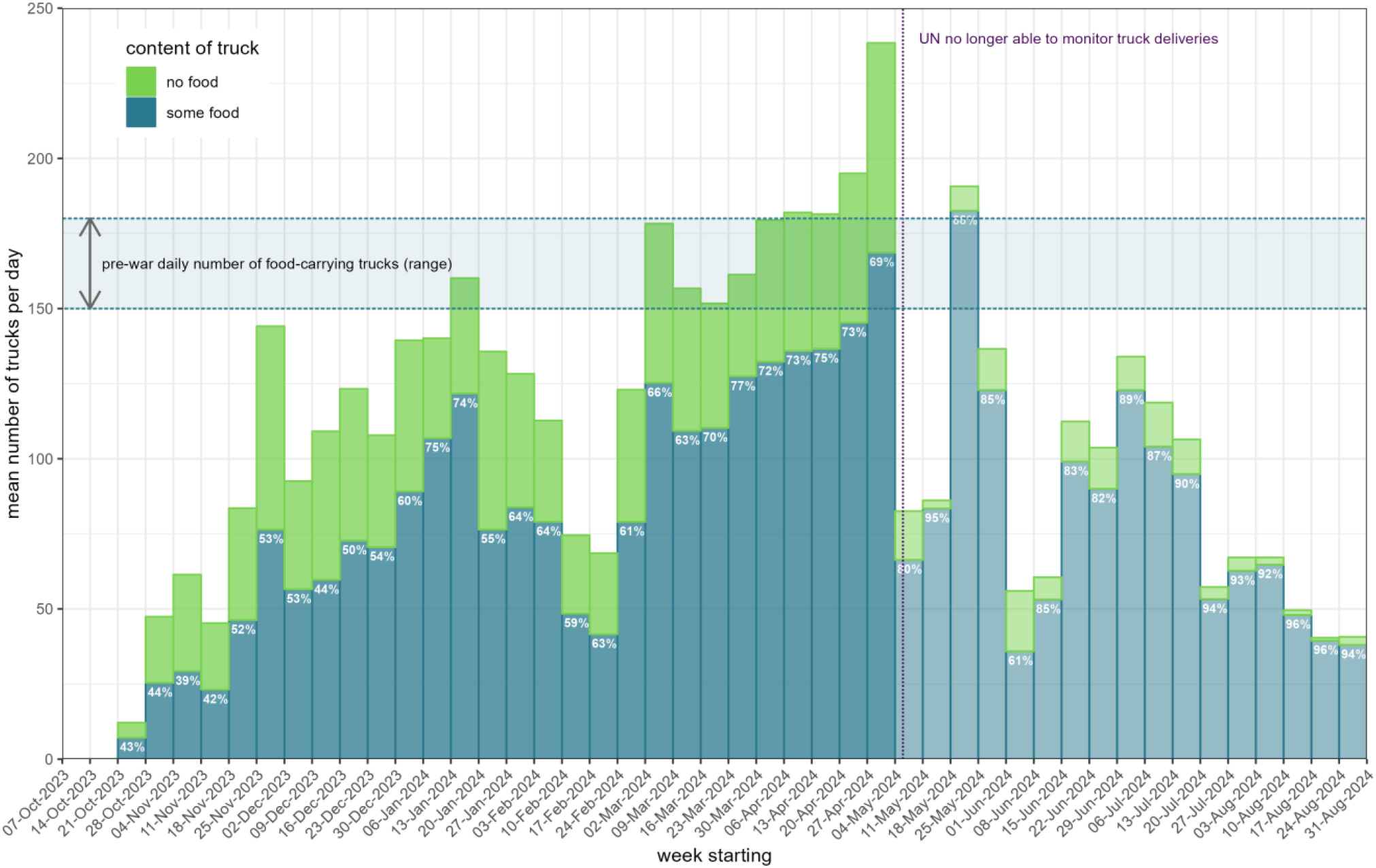
Mean daily number of trucks carrying some food and no food, by week, according to UNRWA data. The pre-war range is as reported by International Phase Classification assessments [22]. Percentages below each bar indicate the proportion of the total weight of items trucked in that consisted of food. Bars are shaded more lightly after the Rafah operation on 5 May 2024, to indicate likely underreporting in the UNRWA dataset.

From mid-November 2023 UNRWA and COGAT yielded widely discrepant trucked-in food weight equivalent, though trends were similar (Figure 2). Over the period when it was directly monitoring all truck arrivals (before 6 May 2024), UNRWA registered 17,032 food-transporting trucks, while COGAT’s database contains 17,924 food records over the same period (it is unclear whether these are all individual trucks). The median (inter-quartile range) estimated weight of food per food-transporting truck was 14.0mt (8.3 to 16.6) per UNRWA, while each COGAT food record featured a median 20mt. Notably, COGAT data show evidence of digit heaping or crude approximation: nearly all food consignment values were reported as exactly 15, 20 or 30mt (Figure 2).

**Figure 2.**
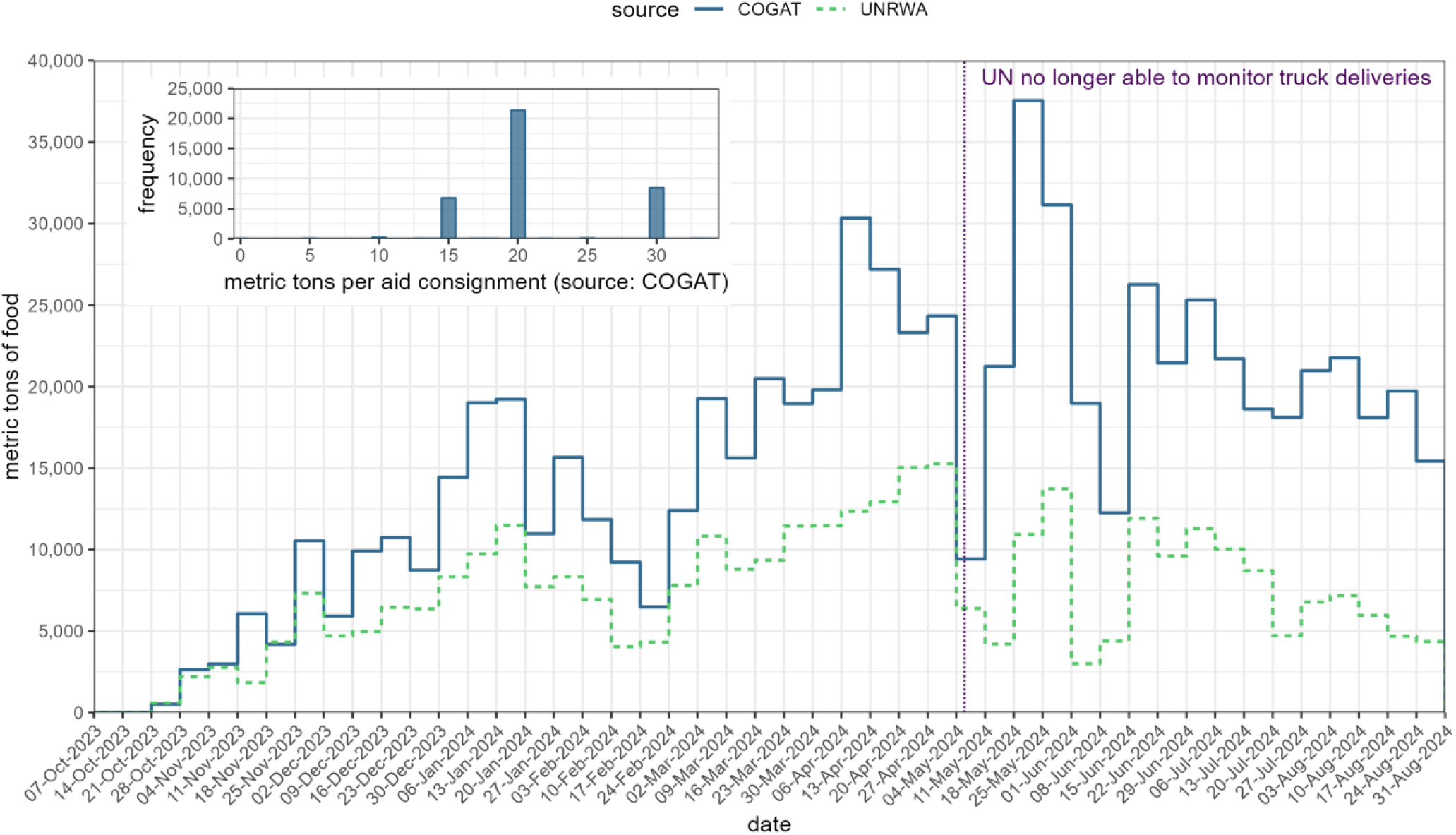
Comparison of food weight equivalent trucked into the Gaza Strip by week, according to UNRWA and the COGAT agency of the Israeli Ministry of Defence. The inset graph shows the distribution of number of metric tonnes per food consignment through any land crossing, according to COGAT data.

### Diversity of food trucked in

A tabulation of individual food items trucked-in and their frequency of importation is provided in the Annex, Table S1. As shown in Figure 3B, a large proportion of food records during the first few months of the war did not have detail on specific foods. After mid-December 2023 around 70-80% of food could be grouped into a food category. Cereals and baked goods accounted for about 40-50% of food weight.

**Figure 3.**
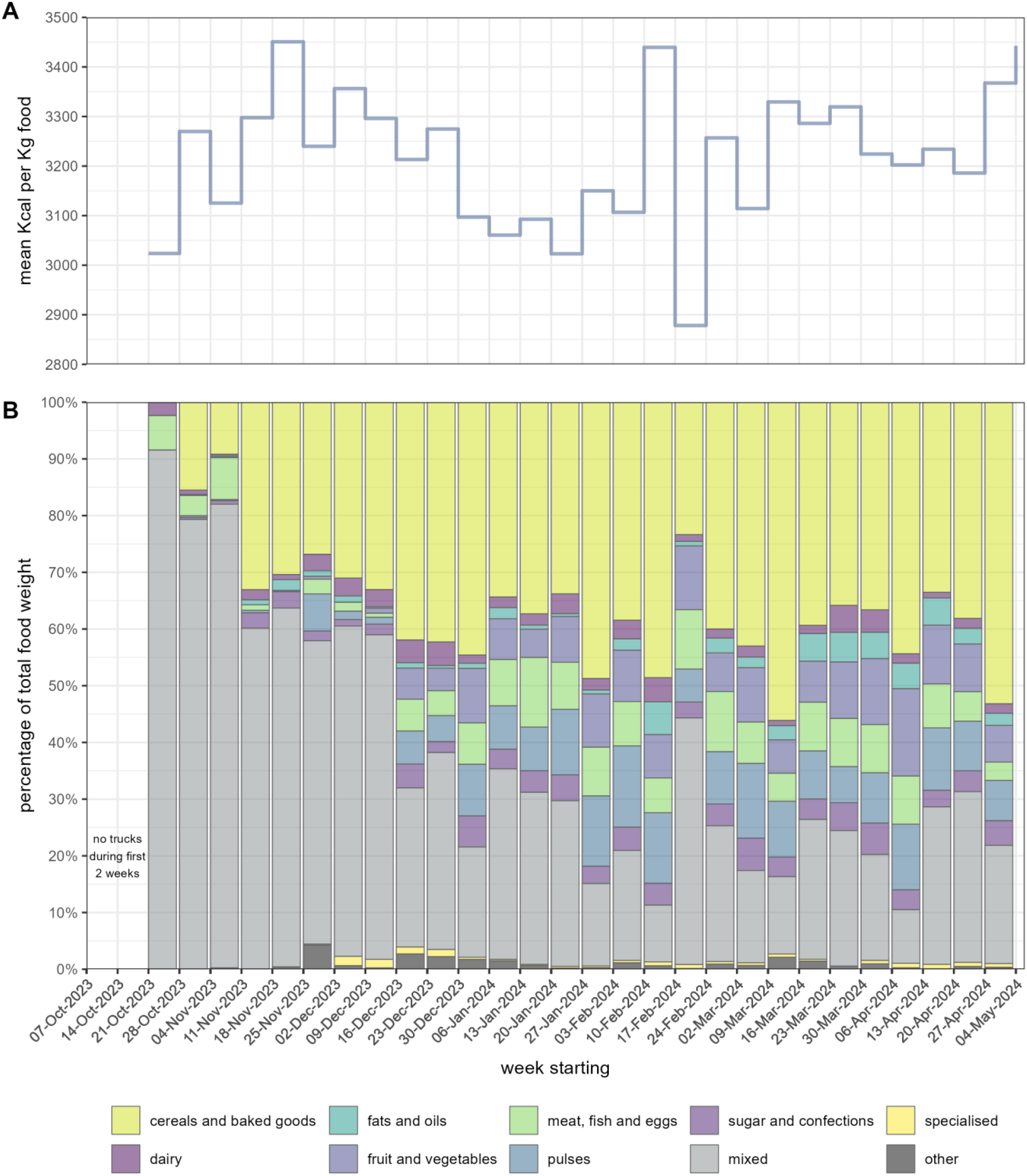
Panel A: estimated mean caloric value of 1 Kg of food trucked into the Gaza Strip, by week. Panel B: proportional contribution of different food categories to the total weight of food trucked in, by week. Only weeks before the Rafah operation are shown. Source: UNRWA dataset. Specialised food includes ready-to-use products, powders and high-nutrient milk formula for the prevention or cure of acute malnutrition.

There was relatively little evolution in terms of the contribution of other food categories. There was, however, considerable fluctuation in the mean caloric value of food (Figure 3A), with the lowest values estimated over the first six weeks of 2024.

### Contribution of different food sources

In south-central Gaza, trucks were the predominant food source, accounting for 75% of total available calories. By contrast, in northern Gaza, trucked-in food contributed only 19% of total calories (Table 1). Conversely, we estimated that stocks present on 7 October 2023 accounted for about half of all Kcal available in the north, but only one fifth in south-central Gaza. The combination of air drops and seaborne deliveries delivered about 3-4% of total calories available in the north and a negligible percentage in south-central Gaza.

**Table 1.**
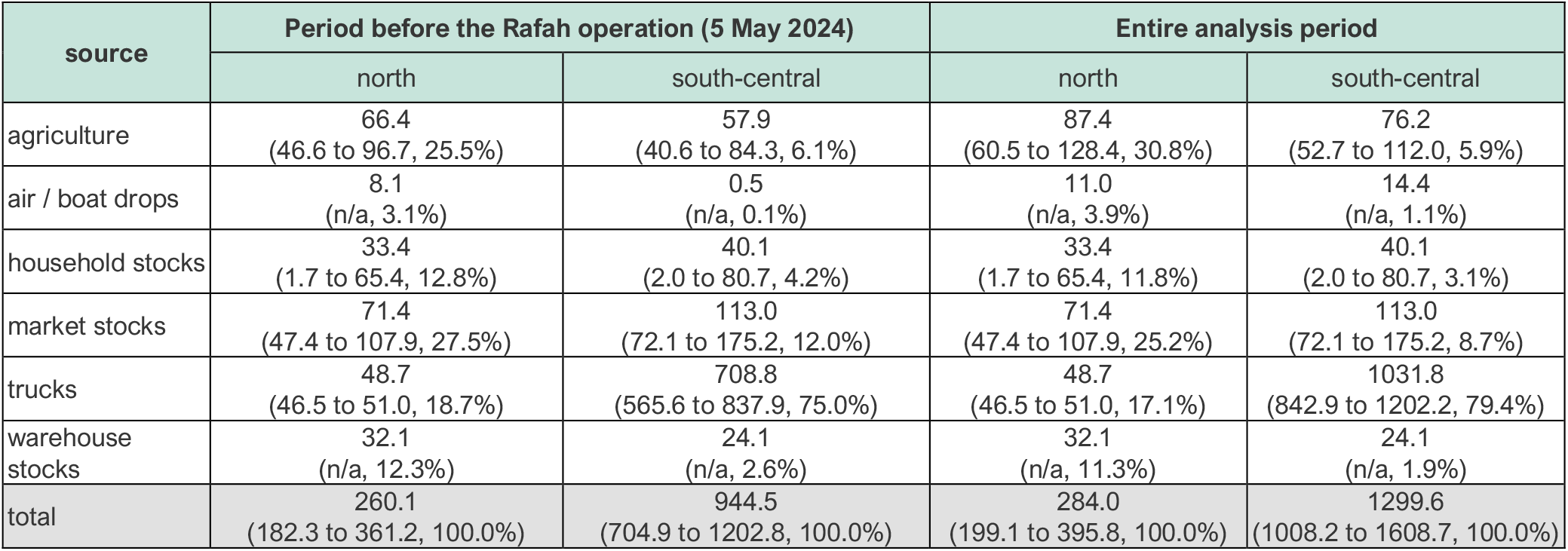
Estimated number of Kcal (in billions) available to the population of northern and south-central Gaza, by period and source. Figures in parentheses report the 95% confidence interval (only shown if the food source featured any uncertainty) and column-wise percentages, relative to the total calories available.

### Per-capita caloric availability

We estimate that the mean number of calories available per person-day began to decline by early November 2023 in the north, where it remained below the recommended daily intake threshold for about 12 weeks (Figure 4), recovering in March 2024 to pre-war baseline levels. In the south-central governorates a more moderate reduction was estimated, with 4 weeks below the recommended intake (note however that we do not account for food losses, distribution problems or inequity in allocation: see Discussion). Estimates for the north are subject to considerable uncertainty, reflecting parameter uncertainty introduced into the simulation.

**Figure 4.**
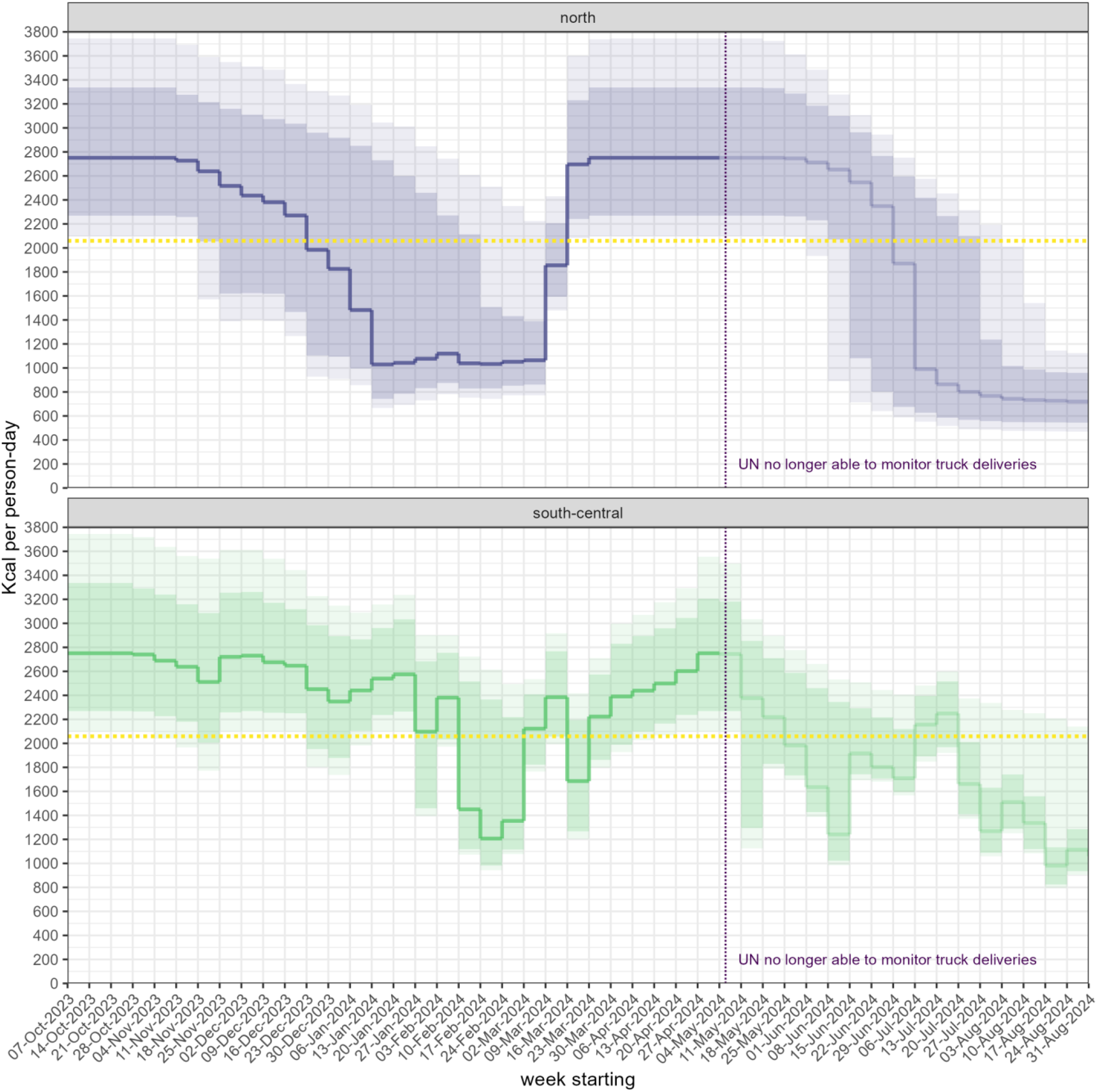
Estimated number of Kcal available per person-day in the north and south-central areas of Gaza, by week. Shaded areas indicate the 80% (darker shade) and 95% (lighter shade) uncertainty interval around the point estimate. The yellow dotted horizontal line denotes the theoretical mean caloric requirement per person-day, based on Gaza’s pre-war demographic characteristics. Point estimates are shaded more lightly after 5 May 2024 to denote data uncertainty and thus likely underestimation following the Rafah operation.

## Discussion

### Main findings

This retrospective analysis finds that the population of Gaza experienced a considerable decline in food availability during the first months of the war. The resulting caloric deficit was probably more severe in the northern governorates, though by then about 85% of Gazans lived in the south-central region. It is plausible that existing stocks at the war’s outset mitigated restrictions on land crossings. The largest source of food remained trucks, and the combination of airdrops and sea deliveries made a proportionately tiny contribution. There was a large discrepancy between UN and Israeli government data. Patterns in the diversity and caloric value of food trucked-in suggest that humanitarian actors may not have optimised the selection of what aid was allowed into Gaza.

There is considerable circumstantial evidence that around the time that we estimate food to have been scarcest in Gaza, food security and nutritional outcomes were indeed deteriorating. The percentage of households reporting a poor food consumption score (<28) had risen from 11% at baseline [10] to 81% in northern governorates and 39% in south-central governorates by November 2023 [22] and 88% and 48-53% respectively by February 2024 [23]. Community middle-upper-arm circumference screenings of children 6 to 23mo in the north reported an acute malnutrition prevalence of 16% in January [46] and 31% in February [47]. However, a steep increase in food availability occurred from late April 2024, coinciding with the reopening of crossings into northern Gaza, and by June acute malnutrition prevalence appeared to be relatively low, despite very limited dietary diversity [47].

Relatedly, our analysis reinforces assessments that high-profile operations to deliver food via air or sea were cost-inefficient and a poor substitute for diplomatic pressure to merely reopen crossings. For example, the 230M USD cost of the JLOTS operation [43] (higher than the entire humanitarian aid budget for the Central African Republic in 2024 [48]) translates to about 37 USD per Kg food or 29 USD per average daily caloric requirement, and fed the equivalent of all of Gaza for ≈4 days based on the recommended intake. Airdrops also accidentally killed at least 21 Gazans [49], though their concentration in the north at the time of greatest need was probably beneficial.

Interestingly, the period of greatest food scarcity was not characterised by obvious changes in the composition of food being trucked-in, and indeed coincided with the lowest caloric value of trucked-in food. Though at any time the majority of food assistance should be familiar to communities and promote healthy, diverse, locally-cooked meals, in a situation of deteriorating food security the distribution of highly nutritious and long-lasting products such as ready-to-use food should also be considered: we saw relatively little such specialised food being imported until May 2024, by which time food availability had improved. The long list of food items that were imported (some of which came through the commercial sector) may suggest insufficient coordination of the food security sector. For example, repeated shipments of croissants, cake and potato crisps (Annex, Table S1) could have been replaced with more appropriate food to support diets, especially among children, pregnant women and the large number of Gazans living with diabetes and/or chronic kidney disease.

### Limitations

The analysis relied heavily on a single UNRWA dataset of truck imports, which appeared highly complete and well-curated, but may be biassed by systematic under- or over-reporting unknown to us. In particular, we computed weight mostly based on the number of pallets transported, but it is possible that pallet load may have varied more than we assumed for particularly heavy or light items, or if truckloads were insufficiently regulated. The UNRWA dataset very probably suffers from underreporting after the IDF’s Rafah operation, and we are accordingly unable to accurately estimate food availability from early May 2024 onwards, though it is noteworthy that both UNRWA and COGAT sources show a worrying decline in food trucked-in after the peak observed in April-May.

We also made numerous assumptions about other food sources, in particular stocks present at the war’s outset and their availability before running out. We attempted to account for uncertainty in these sources through simulation, but it is possible that stocks may have been damaged or been inaccessible to a greater extent than we assumed, and we also did not account for their possible hoarding or selective distribution. There remains considerable uncertainty about population denominators in the north, and even moderate error in these would have affected our Kcal per capita estimates. Gaza’s population has probably decreased due to high mortality and out-migration, though we expect this to have only marginally affected our estimates. Lastly, our analysis provides a crude measure of average caloric availability but does not account for factors such as food loss and inequitable access to food, which may have resulted in lower effective intake overall plus substantial geographic and/or socio-economic group differences in food availability. On balance, therefore, we may have over-estimated food effectively accessible to Gazans.

### Conclusions

Our study adds to increasing evidence on the civilian impacts of the war in Gaza and the conduct of the war. It suggests that Israel, as the *de facto* occupying power, did not ensure that sufficient food was consistently available to the population of Gaza, though the period of resulting caloric deficit was relatively short. In a separate paper we will estimate the resulting effect on nutritional outcomes among Gazan children. It has been suggested that Israel has deliberately starved Gaza’s population [50]; our analysis may support forensic efforts to investigate this. We emphasize that both dietary diversity and caloric quantity are crucial benchmarks for assessing the sufficiency of food provision by occupying powers and humanitarian actors in crisis situations [51].

Situational awareness on food security in Gaza is critically important to inform appropriate humanitarian response. Our analysis indicates that COGAT’s data is not of sufficient quality to guide decision-making. We recommend reinstating UNRWA’s role as an independent and experienced on-the-field monitor of aid and commercial imports.

Lastly, humanitarian actors should review whether there is adequate coordination and technical expertise in place to ensure that what food is allowed into Gaza is both calorically efficient and diverse enough to maintain the best-possible diet, especially for population groups most vulnerable to malnutrition.

## Supporting information

Annex

## Data Availability

All datasets (including URL sources for each datum) and R analysis scripts are available on https://github.com/francescochecchi/gaza_food_availability.

## Acknowledgments

We are grateful to organisations for providing details on the composition of their food parcels, to UNRWA for data on warehouse stocks at the war’s outset and to WFP for data on truck deliveries to the north. This research was conducted with the support of the UK Humanitarian Innovation Hub ([UKHIH](https://www.ukhih.org/)) and the donor, the UK Foreign, Commonwealth & Development Office (FCDO).

## Notes

### Competing Interest Statement

The authors have declared no competing interest.

### Author Declarations

The study was approved by the Ethics Committee of the London School of Hygiene and Tropical Medicine (ref. 29926).

