## Supplementary material for "Wartime food availability in the Gaza Strip, October 2023 to August 2024: a retrospective analysis": Annex

### Caloric equivalent of trucks

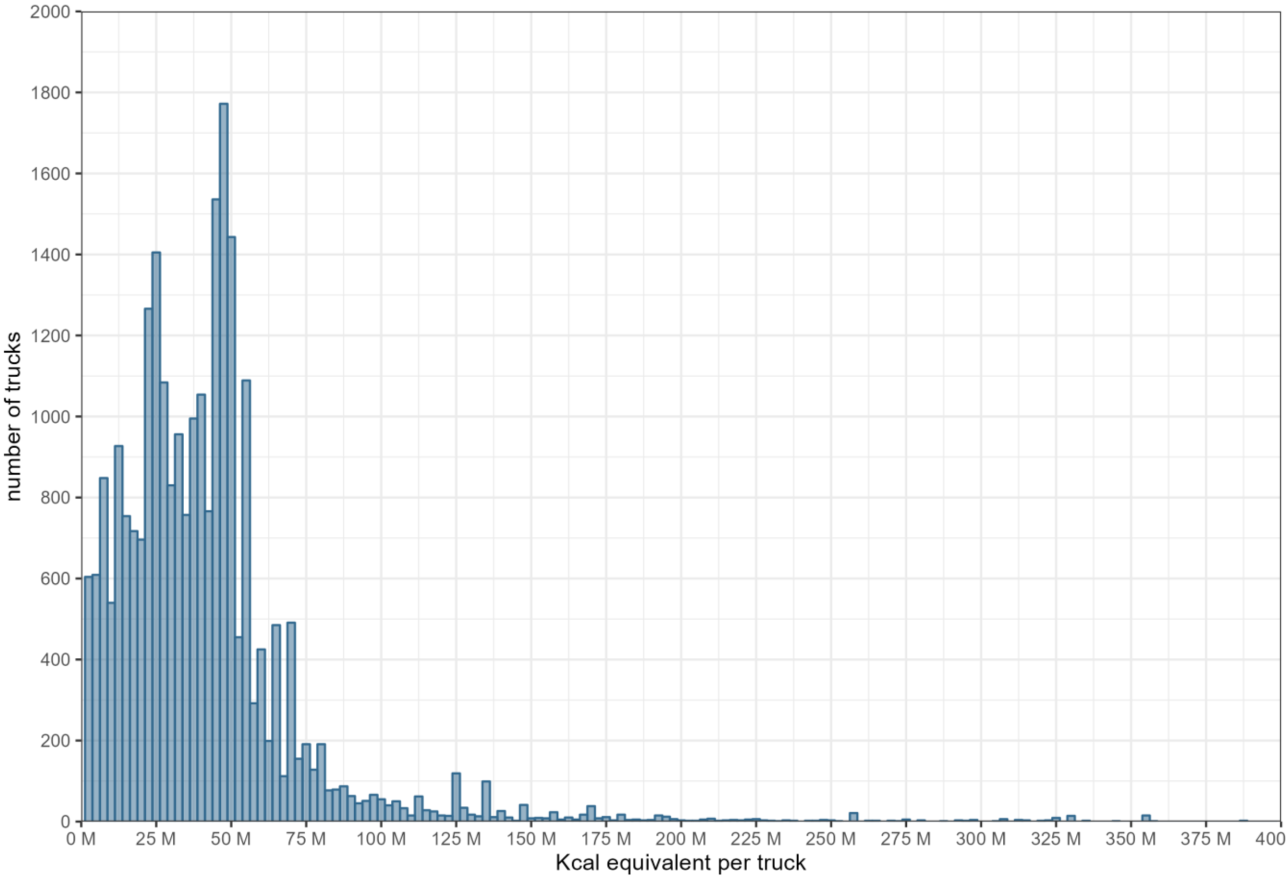


Figure S1. Distribution of estimated Kcal per truck, according to UNRWA data.

### Food items trucked into Gaza

Table S1. Number of times different food items were trucked into Gaza according to the UNRWA database, before and after the 5 May 2024 Rafah operation.

| item | before Rafah operation | post-Rafah operation |
|---|---|---|
| apples | 28 | 0 |
| baby food | 5 | 0 |
| baby milk | 12 | 1 |
| beans | 64 | 3 |
| biscuits | 424 | 290 |
| bread | 17 | 55 |
| bulgur | 2 | 0 |
| cake | 75 | 31 |
| candies | 9 | 32 |
| canned beans | 60 | 11 |
| canned chickpeas | 1 | 0 |
| canned corn | 2 | 4 |
| canned food | 288 | 60 |
| canned hummus | 2 | 0 |
| canned vegetables | 2 | 0 |
| carrot | 1 | 4 |
| cerelac | 3 | 0 |
| cheese | 232 | 52 |
| chicken | 3 | 0 |
| chicken broth | 1 | 0 |
| chickpeas | 24 | 38 |
| chips | 37 | 28 |
| chocolate | 7 | 12 |
| clarified butter | 16 | 40 |
| coconut | 1 | 0 |
| coffee | 38 | 163 |
| cooked food | 1 | 0 |
| cooking oil | 150 | 63 |
| corn | 3 | 10 |
| cream | 1 | 0 |
| croissants | 115 | 0 |
| dairy products | 2 | 0 |
| dates | 141 | 3 |
| dried koshary | 1 | 0 |
| eggplant | 1 | 0 |
| eggs | 53 | 232 |
| fava beans | 3 | 0 |
| fish | 4 | 0 |
| flour | 5575 | 1958 |
| food | 9 | 3 |
| food baskets | 1 | 0 |
| food cartons | 1 | 0 |
| food items | 3326 | 1105 |
| food parcels | 4484 | 962 |
| food supplies | 1 | 0 |
| frozen chicken | 184 | 474 |
| frozen meat | 86 | 174 |
| frozen okra | 5 | 0 |
| frozen vegetables | 7 | 0 |
| fruits | 8 | 1257 |
| garlic | 30 | 3 |
| grapes | 2 | 0 |
| green beans | 1 | 1 |
| halawa | 50 | 2 |
| honey | 22 | 1 |
| hummus | 16 | 3 |
| instant noodles | 36 | 0 |
| jam | 8 | 13 |
| juice | 85 | 410 |
| jute leaves | 2 | 0 |
| kiwi | 1 | 0 |
| legumes | 32 | 69 |

| item | before Rafah operation | post-Rafah operation |
|---|---|---|
| lemons | 19 | 27 |
| lentils | 180 | 89 |
| lentil soup | 1 | 0 |
| luncheon | 178 | 0 |
| meat | 5 | 0 |
| melon | 5 | 1 |
| milk | 155 | 105 |
| noodles | 68 | 367 |
| nuts | 4 | 133 |
| oatmeal | 1 | 0 |
| oil | 246 | 366 |
| onions | 98 | 119 |
| oranges | 37 | 2 |
| pasta | 176 | 31 |
| peanuts | 1 | 0 |
| pears | 1 | 0 |
| peas | 2 | 0 |
| pineapples | 1 | 0 |
| pistachios | 12 | 4 |
| pomegranates | 3 | 0 |
| potatoes | 139 | 91 |
| raisins | 8 | 0 |
| ration packs | 1 | 0 |
| ready meals | 67 | 0 |
| rice | 436 | 200 |
| sage | 9 | 0 |
| salt | 66 | 239 |
| sardine | 1 | 0 |
| sauce | 35 | 35 |
| semolina flour | 25 | 29 |
| sesame seeds | 1 | 0 |
| soda | 23 | 0 |
| spices | 8 | 36 |
| starch | 6 | 24 |
| strawberries | 1 | 0 |
| sugar | 247 | 444 |
| sweet potatoes | 1 | 0 |
| tahini | 4 | 15 |
| tea | 13 | 58 |
| thyme | 2 | 0 |
| tomato paste | 16 | 0 |
| tomato sauce | 61 | 48 |
| tomatoes | 5 | 11 |
| tortilla | 2 | 0 |
| tuna | 84 | 9 |
| vegetable oil | 8 | 0 |
| vegetables | 13 | 716 |
| watermelon | 1 | 24 |
| yeast | 33 | 16 |
| yogurt | 4 | 0 |

### Baseline adult caloric intake

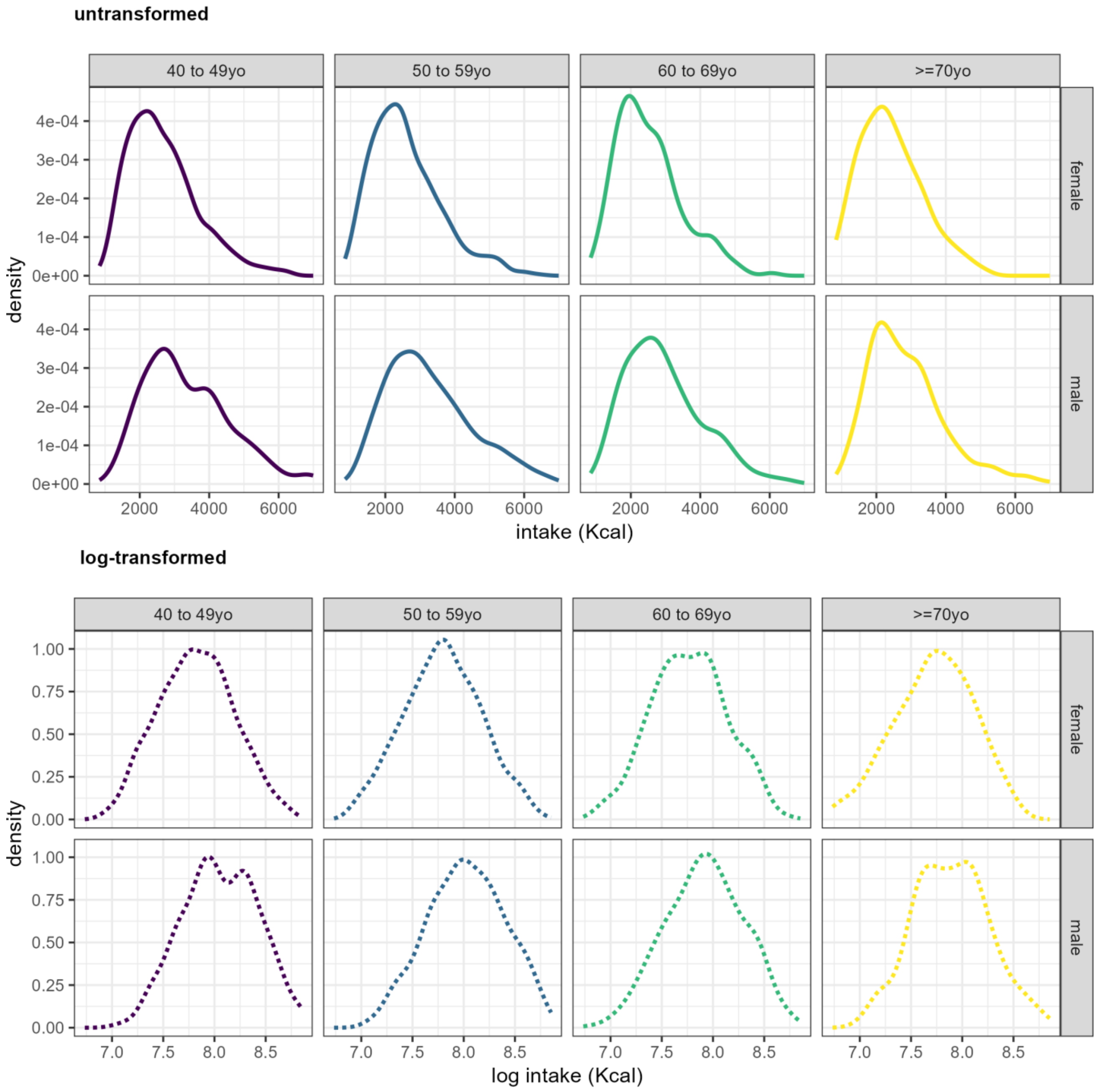


Figure S2. Untransformed and log-transformed distributions of daily caloric intake by age and sex, 2020 [1].

Source:

1. Abu Hamad BA, Jamaluddine Z, Safadi G, Ragi M-E, Ahmad RES, Vamos EP, et al. The hypertension cascade of care in the midst of conflict: the case of the Gaza Strip. J Hum Hypertens. 2023;37:957–68.
